# Glutamate Dehydrogenase as a Superior Biomarker for Choledocholithiasis Risk Stratification

**DOI:** 10.64898/2026.02.14.26346323

**Authors:** Jan P Sutter, Lorenz Kocheise, Soleman Almadok, Jan Drews, Franziska Stallbaum, Jan Kempski, Hanno Ehlken, Hans Pinnschmidt, Marcel Seungsu Woo, Maurice Schückens, Giulia Heide, Lorenz Adlung, Julian Schulze zur Wiesch, Samuel Huber, Ansgar W. Lohse

## Abstract

**Background and Aims:** Choledocholithiasis (CDL) is a common condition that can lead to serious complications, requiring effective risk stratification for timely intervention. While current guidelines use clinical predictors, imaging, and laboratory markers for risk assessment, the role of glutamate dehydrogenase (GLDH) in CDL remains poorly understood. This study aims to evaluate its potential as a clinical biomarker for identifying patients with CDL.

**Methods:** This single-center cohort study identified 23,103 patients who presented to the emergency department of the University Medical Center Hamburg-Eppendorf and underwent routine abdominal laboratory testing between May 2021 and December 2023. Patients were classified into CDL and other diagnoses. To assess the predictive value of age, sex and laboratory markers for CDL, we developed a random forest machine learning model, conducted a backward stepwise logistic regression and performed receiver operating characteristic (ROC) analysis.

**Results:** 152 patients were diagnosed with CDL and 22,951 with other diagnoses. In the random forest machine learning model, GLDH emerged as the most significant feature for predicting CDL. ROC analysis revealed that GLDH had the highest area under the curve of 0.93 among laboratory markers. At the upper limit of normal, GLDH demonstrated the best sensitivity (92%) compared to aspartate aminotransferase (AST), alanine aminotransferase (ALT) and bilirubin. High GLDH levels exceeding 150 U/L demonstrate the highest specificity (99%) for CDL, outperforming AST, ALT and bilirubin.

**Conclusion:** GLDH outperforms AST, ALT and bilirubin as a screening and predictive marker for CDL, supporting its inclusion in clinical guidelines for risk stratification.

## Introduction

Choledocholithiasis (CDL) presents a common clinical challenge due to its potential for severe complications such as cholecystitis, cholangitis, and pancreatitis^1,2^. Risk factors for CDL include advancing age and high-calorie diets^3,4^.

Endoscopic retrograde cholangiopancreatography (ERCP) is performed to remove bile duct obstructions^5^. However, complications such as post-endoscopic sphincterotomy bleeding, perforation, and infection are frequent^6–8^. Therefore, careful patient selection is crucial to minimize complications.

To help risk stratify patients with suspected CDL, the American Society for Gastrointestinal Endoscopy (ASGE) and the European Society of Gastrointestinal Endoscopy (ESGE) have established guidelines based on clinical predictors that include ultrasound imaging of the common bile duct or cross-sectional imaging, clinical symptoms and liver function tests (LFT) including bilirubin and aspartate aminotransferase (AST), alanine aminotransferase (ALT), and alkaline phosphatase (AP)^9,10^. These guidelines stratify patients into low, intermediate, and high-risk categories.

Glutamate dehydrogenase (GLDH) is a mitochondrial enzyme that links carbohydrate and amino acid metabolism and serves as a liver-specific biomarker^11^. It is evenly expressed in the liver lobule ^12^, with lower levels of expression also found in the kidney, pancreas, brain, and intestine^13^. Like ALT, GLDH is released into circulation from damaged hepatocytes when hepatocellular membrane integrity is compromised^14^. Recent preclinical and clinical studies have demonstrated that GLDH serves as a marker of severe hepatocyte injury, potentially exceeding the sensitivity and specificity of traditional transaminases^15–17^. In 1970, a ratio of AST and ALT to GLDH was proposed as an indicator of obstructive jaundice ^18^, but it has not been extensively validated or widely used in clinical practice.

We incorporated GLDH into routine abdominal laboratory testing for patients presenting at the emergency department of the University Medical Center Hamburg-Eppendorf to evaluate its potential as a clinical biomarker for identifying individuals with suspected CDL in this single-center study.

## Methods

### Selection of the study cohort

All patients who presented to the emergency department of the University Medical Center Hamburg-Eppendorf and underwent routine abdominal laboratory testing between May 2021 and December 2023 were identified. All included patients were adults aged 18 years or older. Routine abdominal testing was performed without strict selection criteria, targeting patients with abdominal pain, preexisting abdominal diagnoses, or suspicion of abdominal diseases. Comprehensive data, including sex, age at admission, and laboratory markers (GLDH, AST, ALT, gamma-glutamyltransferase (GGT), AP, bilirubin, C-reactive protein (CRP), and creatinine), along with recent and former diagnoses, were automatically extracted from electronic patient charts. To ensure the correct diagnosis of CDL, we manually reviewed the charts of patients in the automatically generated subgroup with a diagnosis of CDL **(Figure 1)**.

**Figure 1:**
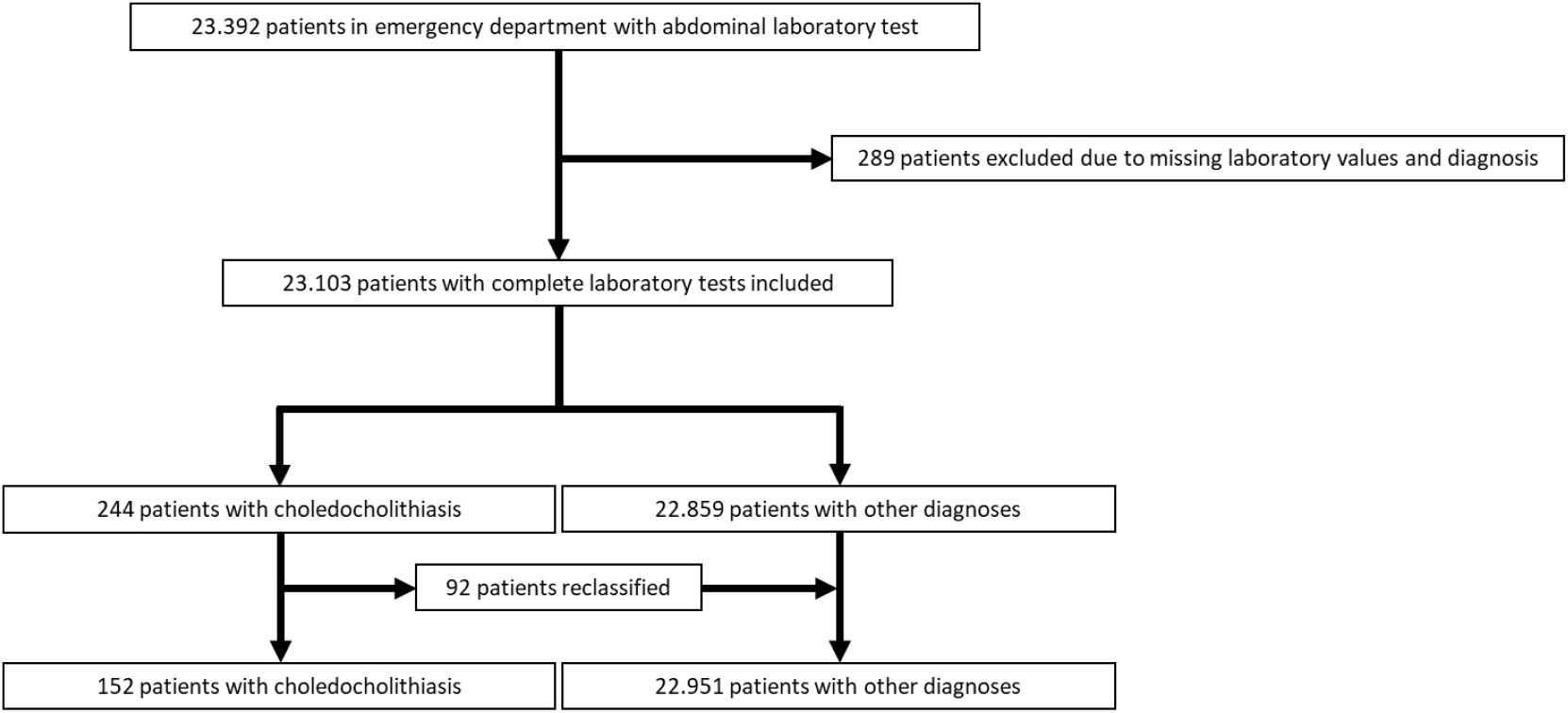
Patient selection: 244 patients with choledocholithiasis were manually reviewed.

The diagnosis of CDL was defined by the detection of intra- and extrahepatic stones via positive imaging (sonography, endoscopic ultrasound, magnetic resonance cholangiopancreatography (MRCP), computed tomography), ERCP, or clinical suspicion of stone passage within 48 hours of laboratory testing. Clinical suspicion of stone passage was diagnosed during the hospital stay, and patient charts were manually reviewed. It was defined by compatible laboratory markers, typical clinical presentation, and spontaneous symptom resolution with negative imaging or ERCP. Patients diagnosed with CDL, with or without cholangitis or cholecystitis, as well as those with acute biliary pancreatitis, were classified under the CDL group, while all other diagnoses were categorized as ‘other’. Patients with missing laboratory markers or missing diagnosis were excluded from the analysis.

### Machine learning

The random forest classifier was trained using an 80:20 train-test split, with regards to equal class distribution. After scaling the data, we used grid search for hyperparameter tuning to identify the best-fitting model for the use case. To address class imbalance, class weight balancing was applied as a constraint for the random forest. This approach was applied to the full set of variables. While the inclusion of bootstrap sampling had minimal impact on the model’s predictive ability, it was ultimately deemed optimal for the dataset, with ‘True’ selected for the bootstrap hyperparameter. The number of features considered for partitioning did not significantly affect model performance, and the value automatically selected by the grid search was used. The optimal hyperparameters for the random forest classifier were identified as follows: {‘criterion’: ‘gini’, ‘max_depth’: 8, ‘max_features’: ‘sqrt’, ‘n_estimators’: 240}.

### Statistical analyses

Continuous variables were analyzed using the Mann-Whitney-U test, while categorical variables were assessed with Fisher’s exact test in two-sided analyses. To assess the influence of independent variables on the probability of CDL, a backward stepwise binary logistic regression analysis was performed. Based on the regression models’ results, we derived a quotient model (GLDH*Bilirubin/AST).

The significant predictors of CDL identified in regression analysis, along with the quotient model, were further evaluated using receiver operating characteristic (ROC) curves. The optimal cutoff value for the quotient model was determined using the Youden Index, and a contingency matrix was generated.

Cohen’s d with 95% confidence intervals were calculated for Figure 3A. Z-scores were generated for each biomarker using the mean and standard deviation of the respective non-CDL groups as references. The z-scores between the biomarkers were compared using unpaired two-sided t-test with FDR multiple comparison correction. Z-scores > 10 were excluded for the graphical representation but were fully included for the statistical tests. Graphs were generated using tidyplots^19^.

A p-value of less than 0.05 was considered statistically significant. All statistical analyses were performed using R (v4.4.1), Python, IBM SPSS Statistics (version 29.0.1.0) and GraphPad Prism (version 10.1.2).

## Results

### Study population

Between May 2021 and December 2023, a total of 23,392 patients underwent routine abdominal laboratory testing in our emergency department. Of these, 289 were excluded due to missing individual laboratory values. Of 244 patients automatically identified with CDL, 92 did not meet the CDL criteria upon manual review and were reclassified into the ‘other’ diagnosis group. The final analysis included 23.103 patients with complete laboratory tests, of whom 152 had CDL and 22.951 had other diagnoses (**Figure 1**).

Patients with CDL (median age 57 years (IQR 43–71)) were significantly older than those with other diagnoses (median age 53 years (IQR 35–69), p = 0.018) (**Table 1**). There were no significant differences in sex distribution or initial serum creatinine levels between the two groups. The median values of all other laboratory markers were significantly higher in the CDL group compared to patients with other diagnoses, including GLDH (109 vs 3 U/L, p < 0.001), AST (197 vs 28 U/L, p < 0.001), ALT (213 vs 23 U/L, p < 0.001), AP (202 vs 82 U/L, p < 0.001), GGT (326 vs 29 U/L, p < 0.001), bilirubin (2.3 vs 0.6 mg/dL, p < 0.001), and CRP (19 vs 8 mg/L, p < 0.001) (**Table 1**).

**Table 1:** Overview of patient characteristics and laboratory markers. Glutamate dehydrogenase (GLDH), aspartate aminotransferase (AST), alanine aminotransferase (ALT), alkaline phosphatase (AP), gamma-glutamyl transferase (GGT). Interquartile range (IQR), number (n).

|  | **Choledocholithiasis (n=152)** | **Other (n=22951)** | **p value** |
| --- | --- | --- | --- |
| **Age (years), median (IQR)** | 57 (43-71) | 53 (35-69) | 0.02 |
| **Sex (male), n (%)** | 87 (57) | 11608 (51) | 0.52 |
| **GLDH (U/l), median (IQR)** | 109 (26-215) | 3 (2-6) | <0.001 |
| **AST (U/l), median (IQR)** | 197 (89-297) | 28 (21- 46) | <0.001 |
| **ALT (U/l), median (IQR)** | 213 (82-364) | 23 (15-39) | <0.001 |
| **AP (U/l), median (IQR)** | 202 (124-329) | 82 (64-113) | <0.001 |
| **GGT (U/l), median (IQR)** | 326 (185-709) | 29 (17-69) | <0.001 |
| **Bilirubin (mg/dl), median (IQR)** | 2.3 (1.2-4.8) | 0.6 (0.4-0.9) | <0.001 |
| **Creatinine (mg/dl), median (IQR)** | 0.9 (0.7-1.1) | 0.9 (0.7-1.2) | 0.92 |
| **C-reactive protein (mg/l), median (IQR)** | 19 (5-61) | 8 (4-38) | <0.001 |

In the CDL group, 115 (76%) patients had imaging or ERCP evidence of stones, while 37 (24%) patients had a clinically high suspicion of stone passage (**Table 2**). Complications occurred in 84 patients (55%), including 50 (33%) with cholangitis and 44 (29%) with biliary pancreatitis.

**Table 2:**
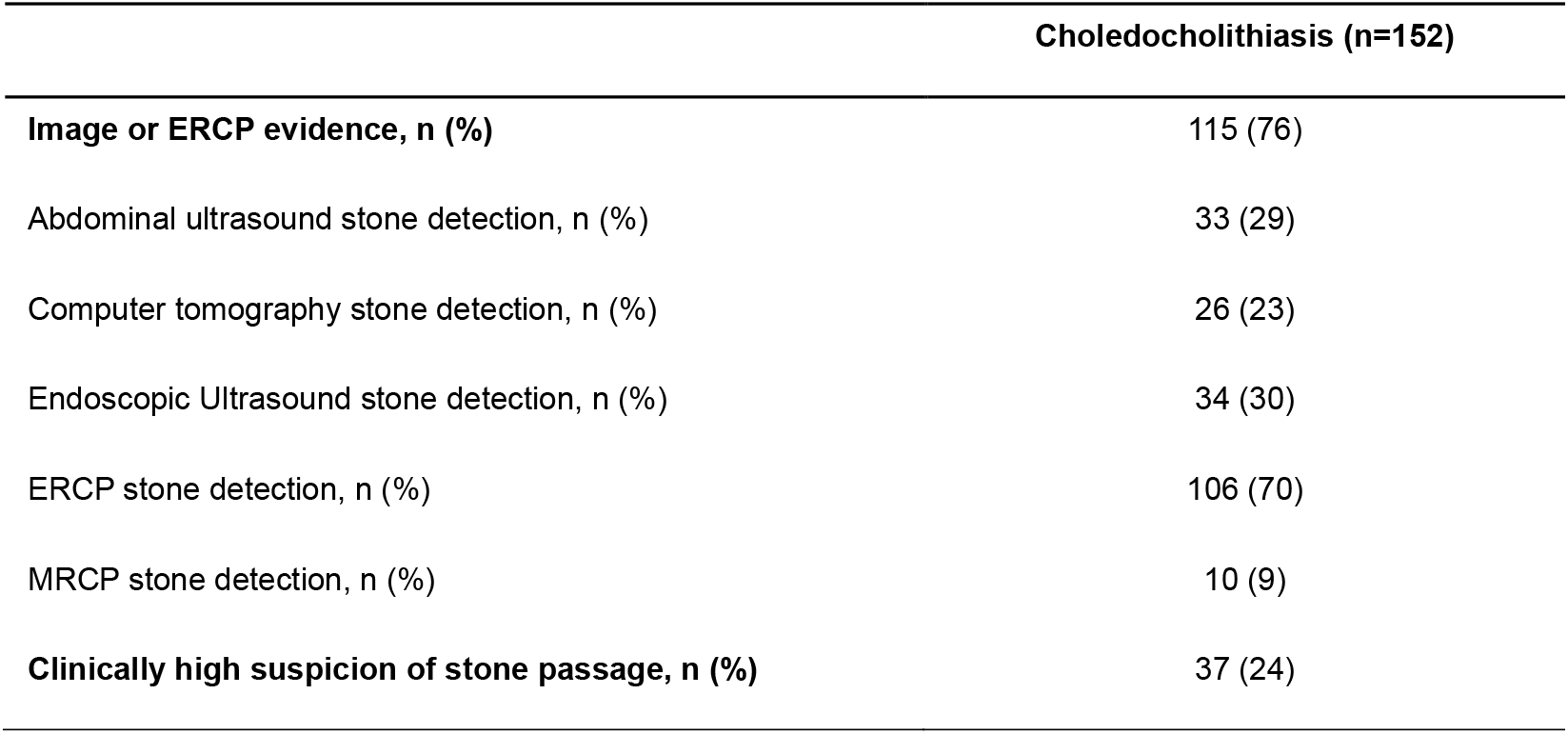
Diagnostic methods and complications of patients with choledocholithiasis. Magnetic resonance cholangiopancreatography (MRCP), endoscopic retrograde cholangiopancreatography (ERCP). Number (n).

|  | **Choledocholithiasis (n=152)** |
| --- | --- |
| **Image or ERCP evidence, n (%)** | 115 (76) |
| Abdominal ultrasound stone detection, n (%) | 33 (29) |
| Computer tomography stone detection, n (%) | 26 (23) |
| Endoscopic Ultrasound stone detection, n (%) | 34 (30) |
| ERCP stone detection, n (%) | 106 (70) |
| MRCP stone detection, n (%) | 10 (9) |
| **Clinically high suspicion of stone passage, n (%)** | 37 (24) |

### Machine Learning – Random Forest

The created random forest demonstrated a good predictive performance and a good fit to the dataset. The optimal number of decision trees in the random forest was found to be 240.

The most relevant features influencing the model’s predictions were GLDH, ALT, bilirubin, AST, and GGT, in descending order of feature importance (**Figure 2A**). The confusion matrix for this model showed a high specificity of 98.4%, sensitivity of 73.3%, negative predictive value (NPV) of 98.4%, positive predictive value (PPV) of 23.2%, and overall accuracy of 98.2% (**Figure 2B**).

**Figure 2:**
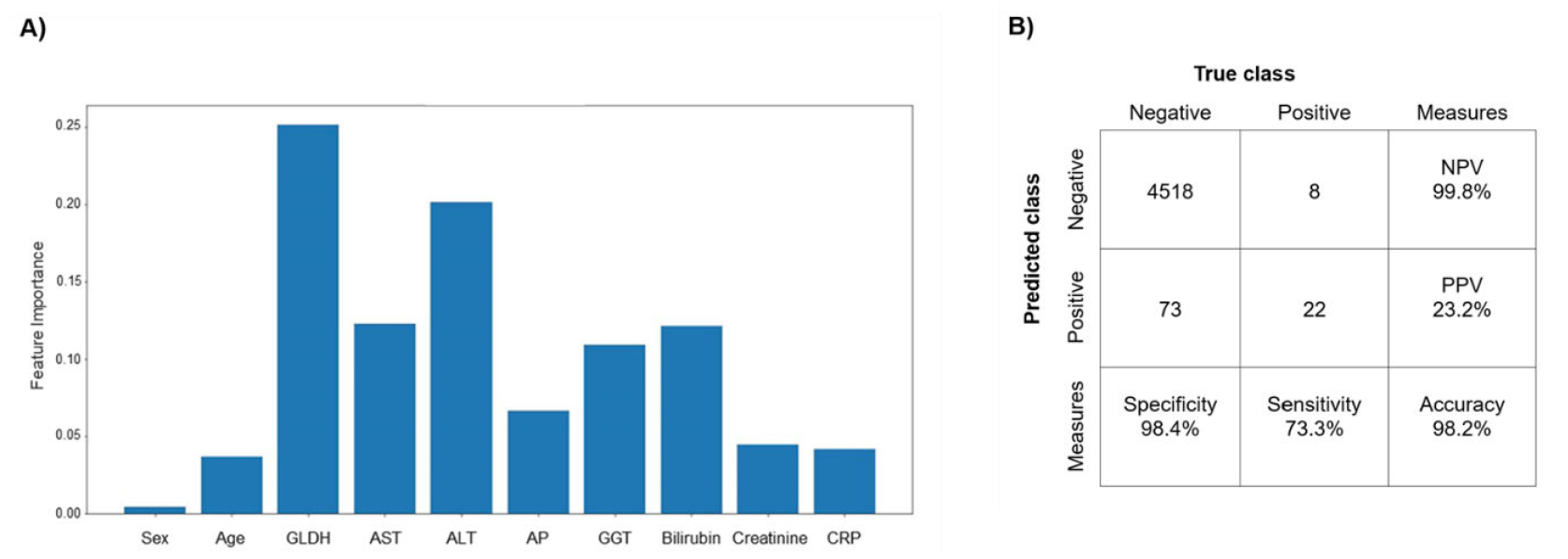
A) Feature importance (%) of sex, age, glutamate dehydrogenase (GLDH), aspartate aminotransferase (AST), alanine aminotransferase (ALT), alkaline phosphatase (AP), gamma-glutamyl transferase (GGT), bilirubin, creatinine, and C-reactive protein in predicting the probability of choledocholithiasis using a random forest model. B) Contingency matrix for the random forest test set, based on the laboratory markers, showing negative predictive value (NPV) and positive predictive value (PPV).

### Predictive Accuracy for Variables

Backward stepwise binary logistic regression indicated that GLDH, ALT, GGT, and bilirubin were associated with an increased likelihood of CDL. The odds ratio (OR) for GLDH was 1.01 (95% CI: 1.00–1.01, p < 0.001) and for bilirubin, it was 1.10 (95% CI: 1.07–1.13, p < 0.001), representing the highest OR. In contrast, AST showed a negative correlation with CDL, with an OR of 0.998 (95% CI: 0.99–1.00, p < 0.001) (**Supplementary table 1**).

In the next step, we tested the predictive accuracies to identify CDL of each biomarker. We first calculated the Cohen’s d effect sizes of the comparison between CDL and the other individuals. In line with our random forest model, GLDH showed the highest effect size, followed by GGT and bilirubin (**Supplementary figure 1**). To directly compare the different biomarkers with each other, we calculated z-scores for the CDL patients using the non-CDL individuals as the reference group (density distributions are shown in **Supplementary figure 2**). Thereby, we detected a significantly stronger difference for GLDH in comparison to all biomarkers except for GGT to separate CDL patients from the non-CDL individuals (**Supplementary figure 3**). Subsequently, we performed ROC curve analysis for the significantly associated variables that showed the following areas under the curve (AUC): GLDH (0.93), AST (0.89), ALT (0.92), GGT (0.90), and bilirubin (0.87), with GLDH exhibiting the numerically highest AUC (**Figure 3A**). After dichotomization using cut-offs, elevated GLDH >7 U/L showed a sensitivity of 92%, higher than ALT >50 U/L (sensitivity: 90%), AST >50 U/L (sensitivity: 88%) and bilirubin >1.1 mg/dL (sensitivity: 78%) (**Table 3**).

**Table 3:** Glutamate dehydrogenase (GLDH), aspartate aminotransferase (AST), alanine aminotransferase (ALT). Specificity, sensitivity, negative predictive value (NPV), and positive predictive value (PPV), each reported in percent (%).

|  | **Specificity** | **Sensitivity** | **NPV** | **PPV** |
| --- | --- | --- | --- | --- |
| **GLDH >7 U/l** | 78 | 92 | 99.9 | 3 |
| **GLDH >150 U/l** | 99 | 41 | 99.6 | 25 |
| **ALT >50 U/l** | 81 | 90 | 99.9 | 3 |
| **ALT >320 U/l** | 97 | 35 | 99.6 | 10 |
| **AST >50 U/l** | 78 | 88 | 99.9 | 3 |
| **AST >260 U/l** | 97 | 35 | 99.6 | 7 |
| **Bilirubin >1.1 ml/dl** | 82 | 78 | 82 | 3 |
| **Bilirubin >3.5 ml/dl** | 96 | 35 | 99.6 | 6 |

At concentrations above 150 U/L, GLDH reached its highest PPV of 25%, with a sensitivity of 41% and a specificity of 99%, which was higher than the highest PPVs observed for ALT > 320 U/L (10%; sensitivity 35%, specificity 97%), AST > 260 U/L (7%; sensitivity 35%, specificity 97%), and bilirubin > 3.5 mg/dL (6%; sensitivity 35%, specificity 96%) (**Table 3**).

### CDL quotient model

Based on the back stepwise binary logistic regression, we derived a quotient model for CDL outcome as follows: GLDH * Bilirubin / AST. The variables in this quotient model reflect the statistically significant associations and their directionality, as revealed in the regression analysis.

The ROC curve for the quotient model demonstrated an AUC of 0.95, higher than all individual laboratory markers, with the maximum Youden index at 0.26 (**Figure 3B**). The contingency matrix for the optimal quotient model of >0.26 showed specificity of 90%, sensitivity of 92%, NPV of 99.9%, PPV of 5.5% and accuracy of 89.5% (**Figure 3C**).

**Figure 3:**
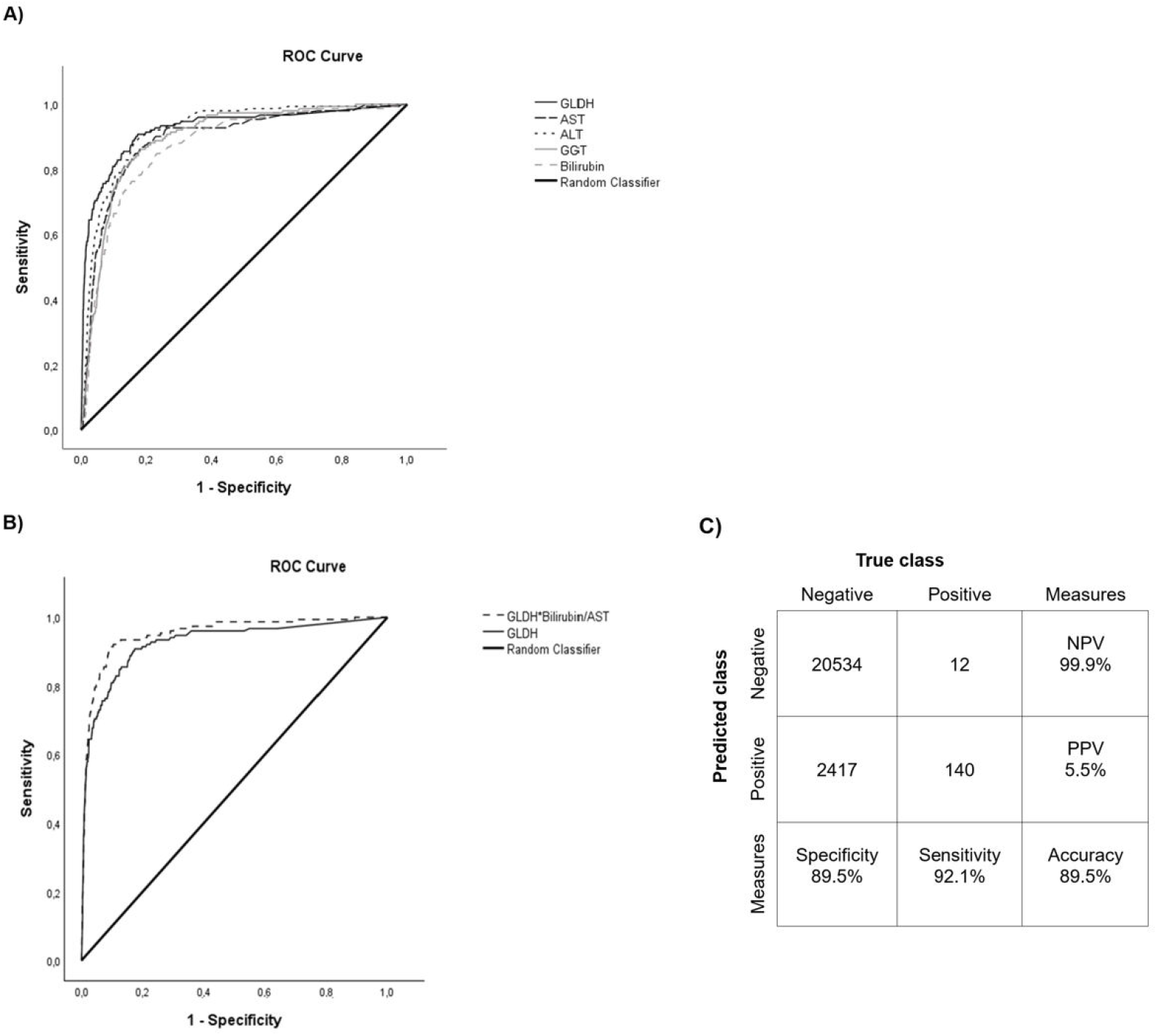
(A) Receiver operating characteristic (ROC) curves for laboratory markers, area under the curve (AUC): GLDH: 0.93, AST: 0.89, ALT: 0.92, bilirubin: 0.87, GGT: 0.90, Youden index maximum: GLDH 8,95 U/l, AST: 56,5 U/l, ALT 51,5 U/l, GGT: 100,5 U/l, bilirubin: 2.0 mg/dl. (B) Receiver operating characteristic (ROC) curves for GLDH and quotient, area under the curve (AUC): Quotient: 0.95, GLDH: 0.93; Youden index maximum: Quotient 0.26, (C) Contingency matrix and calculated measures for Quotient >0.26. Glutamate dehydrogenase (GLDH), aspartate aminotransferase (AST), alanine aminotransferase (ALT), alkaline phosphatase (AP), gamma-glutamyl transferase (GGT), negative predictive value (NPV), positive predictive value (PPV).

## Discussion

This study aimed to evaluate the role of GLDH as a clinical biomarker to identify patients with CDL presenting to the emergency department. GLDH demonstrated the strongest discriminative and predictive performance of all available biomarkers, making it the most accurate marker for identifying patients with CDL in our cohort.

Notably, GLDH exhibited the highest feature importance for prediction in our random forest model, surpassing well-established markers such as AST, ALT, AP, and bilirubin which are used in the ASGE and ESGE guidelines ^9,20–22^. The random forest classifier exhibited solid performance, achieving both a high PPV and good sensitivity, despite the relatively unrestricted patient selection. Applying machine learning models to more refined datasets, may further improve decision-making in patients with suspected CDL.

In a meta-analysis of CDL predictors, seven studies using abnormal LFTs reported a mean sensitivity of 85% (range: 71–99%)^23^, which is consistent with our findings. The sensitivity of 92% for GLDH compares favourably and exceeds the sensitivity of LFTs and bilirubin in our cohort. This highlights its potential role as a screening tool in future guideline recommendations.

When aiming for the highest PPV, GLDH demonstrated an even greater benefit compared to other LFTs and bilirubin. As our cohort was solely selected based on routine abdominal laboratory testing, we hypothesize that in a more defined patient group, including clinical symptoms and physical examination, the PPV of markedly elevated GLDH would be even higher. According to the ASGE guidelines, patients with a dilated bile duct or elevated abnormal liver biochemical tests or age over 55 years in the absence of stones on imaging, with symptomatic cholelithiasis are classified as at intermediate risk for choledocholithiasis. For this group, an EUS or MRCP prior to a potentially unnecessary ERCP is recommended^9^. We postulate that additional information from GLDH, with its high PPV, could aid in risk stratification and potentially reduce the number of EUS examinations required in this patient population. This could not only optimize diagnostic pathways but also lower healthcare costs and reduce the burden of invasive diagnostic procedures for patients.

In our cohort, median GLDH levels were elevated to approximately 16 times the upper limit of normal (ULN, 7 U/L), compared to a roughly 4-fold increase seen in ALT or AST (ULN, 50 U/L). Studies suggest that GLDH is more liver-specific than conventional LFTs, which could explain its markedly higher increase and stronger diagnostic value in our study^15–17,24^. We hypothesize that this marked rise in GLDH may result from direct acute trauma caused by a stone, leading to a disproportionately higher elevation in GLDH compared to conventional LFTs. Further advantages of GLDH over ALT may lie in its shorter half-life (16-18 hours vs. 47 hours), making it a more responsive marker for acute liver damage^12,25^. This rapid clearance allows GLDH to reflect ongoing liver injury more accurately and may provide insights into the progression of hepatic damage, which other LFTs may not capture as effectively. Additionally, this shorter half-life matches our clinical observation that GLDH levels decline rapidly after a potential spontaneous stone passage, especially when compared to other liver function tests. While the timing of ERCP in patients with cholangitis is well defined in the Tokyo 2018 guidelines^26^, the optimal timing in patients without or with mild cholangitis remains less well defined and is usually elective. In this setting, monitoring GLDH dynamics before an elective ERCP could help identify a potential spontaneous stone passage and thereby avoid unnecessary interventions.

To enhance clinical utility, we developed the CDL quotient model based on our regression analysis. ALT was excluded from the model, as its inclusion did not yield additional predictive value. This may be explained by the strong correlation between GLDH and elevated ALT levels reported by Zelder and colleagues^14^. Interestingly, the quotient model achieved a higher AUC than any single marker. However, GLDH alone outperformed the quotient in terms of optimal PPV and sensitivity, key parameters for risk stratification in clinical guidelines.

Notably, Zelder and colleagues postulated a GLDH-based quotient to be predictive of obstructive jaundice and gallstone disorders in 1970^18^. However, this quotient has not been further evaluated and not been widely adopted in clinical practice, possibly because commercially available GLDH reagents only became broadly accessible in the early 2000s^27^.

We hypothesize that GLDH can also serve as a valuable marker for differentiating between biliary and non-biliary pancreatitis. An ALT value exceeding three times the upper limit of normal within the first 48 hours is considered highly suggestive of gallstone-related pancreatitis^25^. Given the superior predictive power of GLDH observed in our study, its diagnostic potential should be further investigated.

Despite the robust performance of GLDH, this study has several limitations. First, it is a single-center analysis, which limits the generalizability of the results to other populations or healthcare settings. Additionally, while this study focused on laboratory markers, other clinical factors such as comorbidities or concurrent medications were not fully explored and could influence the interpretation of the results. However, the baseline characteristics of our study population align with findings from previous studies on CDL, particularly in terms of age and laboratory values^28^. Additionally, the ratio of patients with positive imaging evidence for CDL and those with a clinically high suspicion of stone passage is similar to reports in other studies^28–30^.

## Conclusion

Our study highlights the superior diagnostic performance of GLDH as a screening and predictive laboratory marker for CDL compared to established LFTs. These results support the inclusion of GLDH in current clinical guidelines for the risk stratification of patients with suspected CDL.

## Data Availability

All data produced in the present study are available upon reasonable request to the authors

## Statements and Declarations

### Competing interests

The authors declare no competing interests.

### Consent to participate and consent to publish

We received a waiver for consent to participate and consent to publish from the Ethics Committee of the Hamburg Medical Association since we used only anonymized data (Application No. 408062-WF).

### Grant Support

Funding Jan Peter Sutter: Funded by the Deutsche Forschungsgemeinschaft (DFG, German Research Foundation) – Project number 493624519.

## Notes

### Competing Interest Statement

The authors have declared no competing interest.

