## Supplemental material for "Glutamate Dehydrogenase as a Superior Biomarker for Choledocholithiasis Risk Stratification"

### Supplementary Materials

#### Supplementary figure 1

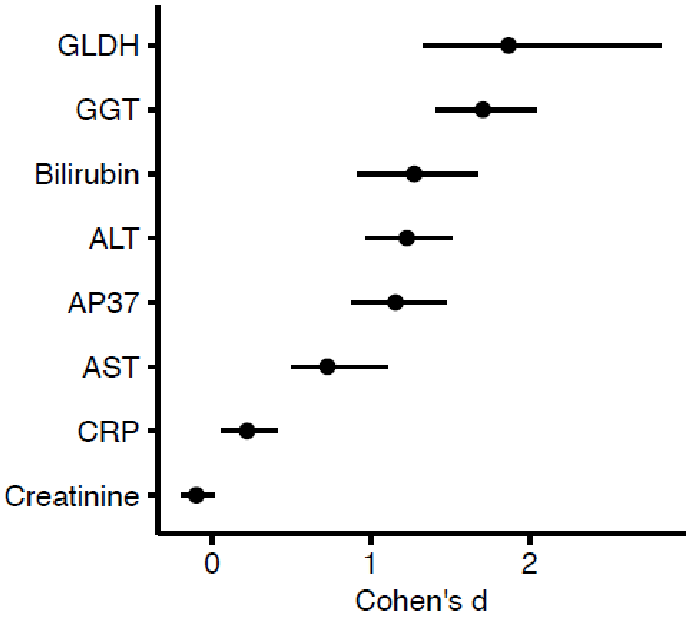


Supplementary figure 1: Cohen's d with 95% confidence intervals of the statistical comparisons of the indicated biomarkers between CDL patients and non-CDL individuals. The biomarkers are sorted by the effect size. Glutamate dehydrogenase (GLDH), aspartate aminotransferase (AST), alanine aminotransferase (ALT), alkaline phosphatase (AP), gamma-glutamyl transferase (GGT).

#### Supplementary figure 2

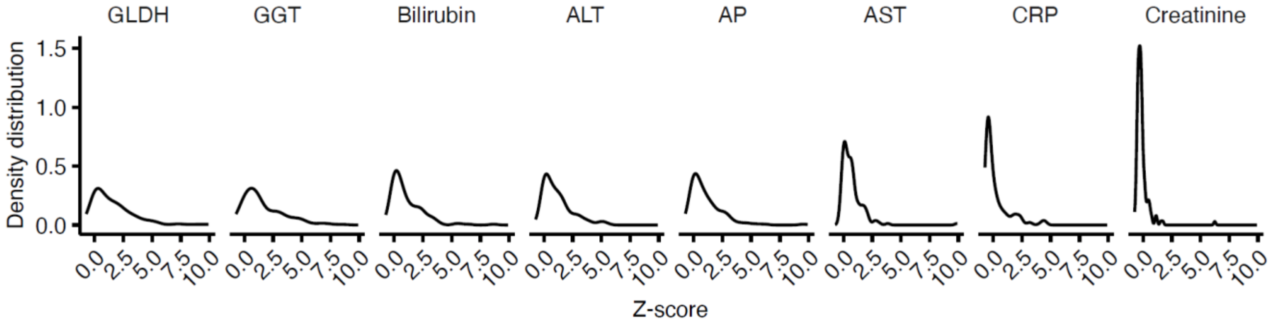


Supplementary figure 2: Density plots of the Z-scores of the indicated biomarkers were generated using the non-CDL individuals as reference group. The biomarkers are sorted by the effect size determined in supplemental figure 2. Glutamate dehydrogenase (GLDH), aspartate aminotransferase (AST), alanine aminotransferase (ALT), alkaline phosphatase (AP), gamma-glutamyl transferase (GGT).

### Supplementary figure 3

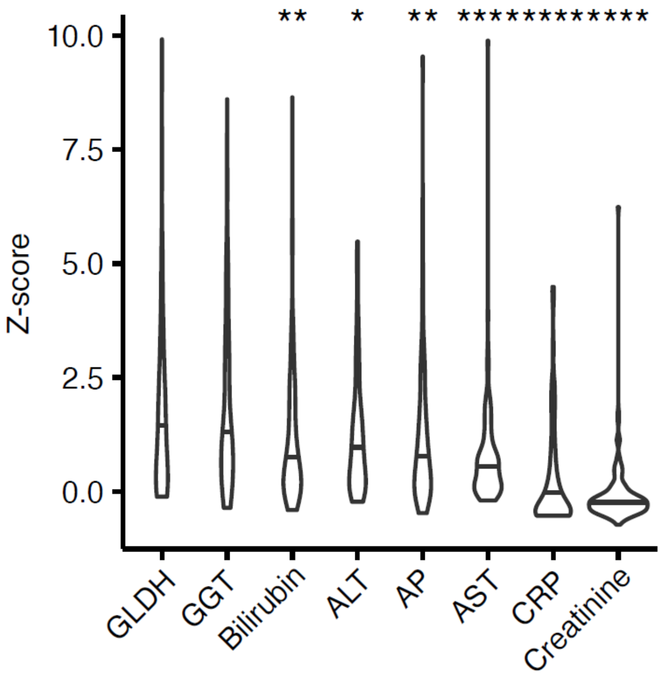


Supplementary figure 3: Z-scores of the indicated biomarkers were generated using the non-CDL individuals as reference group. Statistical comparisons between GLDH and all other biomarkers were performed using FDR-corrected unpaired t-tests. The biomarkers are sorted by the effect size determined in supplemental figure 2. *p < 0.05, **p < 0.01, ****p < 0.0001. Glutamate dehydrogenase (GLDH), aspartate aminotransferase (AST), alanine aminotransferase (ALT), alkaline phosphatase (AP), gamma-glutamyl transferase (GGT).

### Supplementary table 1

| | Initial Model | | | Final Model | | |
|---|---|---|---|---|---|---|
| | OR | 95%CI for OR | p-value | OR | 95%CI for OR | p-value |
| **GLDH** | 1.01 | 1.00-1.01 | <0.001 | 1.01 | 1.00-1.01 | <0.001 |
| **AST** | 1.00 | 0.99-1.00 | <0.001 | | 0.99-1.00 | <0.001 |
| **ALT** | 1.00 | 1.00-1.00 | <0.001 | 1.00 | 1.00-1.00 | <0.001 |
| **AP** | 1.00 | 1.00-1.00 | 0.624 | | | |
| **GGT** | 1.00 | 1.00-1.00 | <0.001 | 1.00 | 1.00-1.00 | <0.001 |
| **Bilirubin** | 1.10 | 1.01-1.13 | <0.001 | 1.01 | 1.07-1.130 | <0.001 |
| **Creatinine** | 0.78 | 0.61-1.02 | 0.065 | | | |
| **C-reactive Protein** | 1.00 | 1.00-1.00 | 0.218 | | | |
| **Sex** | 1.11 | 0.77-1.55 | 0.615 | | | |
| **Age** | 1.01 | 1.00-1.02 | 0.052 | | | |
| **Constant** | 0.01 | | <0.001 | 0.01 | | <0.001 |

Supplementary table 1: Backward stepwise binary logistic regression: Impact of glutamate dehydrogenase (GLDH), aspartate aminotransferase (AST), alanine aminotransferase (ALT), alkaline phosphatase (AP), gamma-glutamyl transferase (GGT), bilirubin, creatinine, C-reactive protein, sex and age on the probability of choledocholithiasis. Odds ratio (OR), confidence interval (CI).
